# Can local farms sustain Thai school lunches? A qualitative exploration of challenges and solutions

**DOI:** 10.1101/2025.10.14.25338034

**Authors:** Panrawee Praditsorn, Tina Beuchelt, Christian Borgemeister, Ute Nöthlings, Nuttarat Srisangwan, Arisa Keeratichamroen, Kitti Sranacharoenpong

## Abstract

School lunch programs play a critical role in providing essential nutrition to children; however, sourcing ingredients locally remains a challenge for many schools in Thailand. This study aimed to explore the drivers and barriers influencing the integration of local farms, using Chiang Mai as a case study, into the Thai school lunch program. In-depth, semi-structured interviews were conducted with 13 school staff and six local farmers in Saraphi and Hang Dong districts of the Chiang Mai area. Data were initially analyzed thematically to identify key patterns and concepts. These themes were then examined using a SWOT (Strengths, Weaknesses, Opportunities, Threats) framework to categorize internal and external factors influencing local food sourcing. A TOWS (Threats, Opportunities, Weaknesses, Strengths) matrix was used to develop context-specific strategies for enhancing school-farm collaboration. The findings showed that key drivers for school-farm integration included strong community support, local market access, and agricultural education initiatives. Barriers included seasonal variability, inconsistent supply, and strict budgetary constraints. The study provided practical recommendations to enhance the sustainability of school-farm integration, including improved procurement planning, streamlined administrative processes, and the development of supportive policies to facilitate farm-to-school partnerships.

## Introduction

Child nutrition is fundamental for optimal physical and cognitive development, impacting academic performance, health outcomes, and overall well-being [1]. School meal programs play a crucial role in ensuring children receive essential nutrients [2]. In Thailand, the national school lunch program serves as a vital safety net, providing daily nutrition to students in public primary schools who rely on these meals [3]. However, ensuring the effectiveness of this program necessitates a reliable supply of high-quality food, posing a challenge for many schools.

To address this challenge, local food sourcing, also known as “farm-to-school” initiatives, has emerged as a promising strategy. Research suggests that sourcing locally provides fresher produce, which retains higher nutritional value due to reduced transportation time and minimal nutrient degradation [4, 5]. Additionally, it supports the local economy [6], reduces transportation-related carbon emissions [7], and often involves the use of fewer synthetic pesticides, resulting in safer and healthier food options for children [8, 9].

Studies highlight the growing adoption of local food sourcing in school meal programs. In the United States, Jefferson Elementary School in Riverside, California, partnered with the Center for Food and Justice to establish a farm-to-school salad bar program [10], while the Fresh from the Farm initiative in Chicago provided locally sourced meals and educational opportunities in selected schools [11]. A more recent study in North Carolina on Farm to Early Care and Education identified strategies used by childcare centers to source local food, including partnerships with distributors and retailers, as well as joint purchasing with other centers and families [12]. In Asia, Japan’s national Shokuiku (food education) policy has supported community-based school lunch programs that connect schools with local farmers, integrating nutrition and agricultural education [13]. Similarly, South Korea has introduced eco-friendly food policies that promote local sourcing through direct farmer contracts and government subsidies, ensuring affordability [14]. In Thailand, although empirical research remains limited, a study reported that school vegetable gardens promote vegetable consumption among children [15]; however, limited space restricts the implementation of such initiatives in many schools. More recently, strengthening school lunch management in Thailand has been proposed by fostering collaboration among school staff and expanding partnerships with external communities [16].

Despite the recognized benefits of local sourcing, widespread adoption in school lunch programs faces obstacles. These include challenges related to inconsistent supply and seasonal variability, transportation difficulties in remote schools, and natural disasters such as flooding, which impact the quality of ingredients [16]. Additional challenges involve potentially higher costs compared to conventional procurement methods [17, 18] and logistical constraints related to storage, distribution, and processing [19]. While these obstacles have been documented in various settings, there remains a limited understanding of the specific drivers and barriers affecting the implementation of local sourcing within the Thai school lunch context. This knowledge gap impedes the development of effective, contextually appropriate strategies to foster integration between Thai schools and local farmers.

This study aimed to address this gap by investigating the drivers and barriers experienced by local farmers and school staff in establishing partnerships within the Thai school lunch program. It was guided by the following key questions: (1) What are the drivers (strengths and opportunities) and barriers (weaknesses and threats) that influence integration between the Thai school lunch program and local farms? and (2) What strategies can enhance the integration between Thai school lunch programs and local farms?

To address these questions, a qualitative research design was employed, incorporating SWOT-TOWS (Strengths, Weaknesses, Opportunities, Threats - Threats, Opportunities, Weaknesses, Strengths) analysis. This framework enabled the systematic identification of key internal and external factors affecting school-farm collaboration. Through this lens, the study developed actionable, context-specific strategies to facilitate the integration of local sourcing into the Thai school lunch program.

## Method

### Study design

This study was a part of a larger research project of a nutrition education intervention program in Chiang Mai, Thailand. A qualitative research method was employed to explore the perspectives of school staff responsible for managing the lunch program and local farmers in areas surrounding the schools regarding school-farm integration in the Chiang Mai area.

### Participants’ recruitment

Ten public primary schools, five from each of the Saraphi and Hang Dong districts in Chiang Mai, were randomly selected as a part of the larger research project. The school selection process used a multistage sampling design, where both districts were chosen due to their similarities, including a semi-urban context, comparable population densities, demographic and socioeconomic profiles, and their inclusion in the same educational service area. These similarities allowed for the combination of interview data from both districts to create a comprehensive understanding of the questions under study.

In each school, one to two staff members responsible for school lunch management were selected for face-to-face, in-depth interviews, as they possessed detailed information about the daily ingredients and their sources. Information on local farm locations was also obtained from these staff members. If they sourced ingredients from local farms, they were asked to provide farm locations, and visits to these farms were arranged with their help. If the staff did not purchase from local farms, they were asked if they knew the locations of any nearby farms, leading to visits organized with their assistance.

Road network distance analysis was conducted using QGIS 3.38.3 (Grenoble) to calculate the shortest travel distances in kilometers (km) between schools, markets, and local farms. The shortest path (point-to-point) tool was used to compute distances along the actual road network, rather than relying on straight-line (Euclidean) measurements. This approach provided a more realistic representation of geographic accessibility in the study area.

Fig 1 illustrates the locations of schools, markets where school staff purchased ingredients for school lunches, and local farms. Farmer 1 was recommended by a local contact who was working with local stakeholders at the time of data collection and assisted in facilitating initial contacts. Farmers 2 and 3 were suggested by the teacher from School 5, and Farmer 4 was referred by the director of School 10. Farmers 5 and 6 were recommended by the director of School 6. Although Farmer 7 was also recommended by the cook of School 4, he was unavailable during the data collection period despite multiple attempts by the researcher and the cook to arrange an interview.

**Fig 1.**
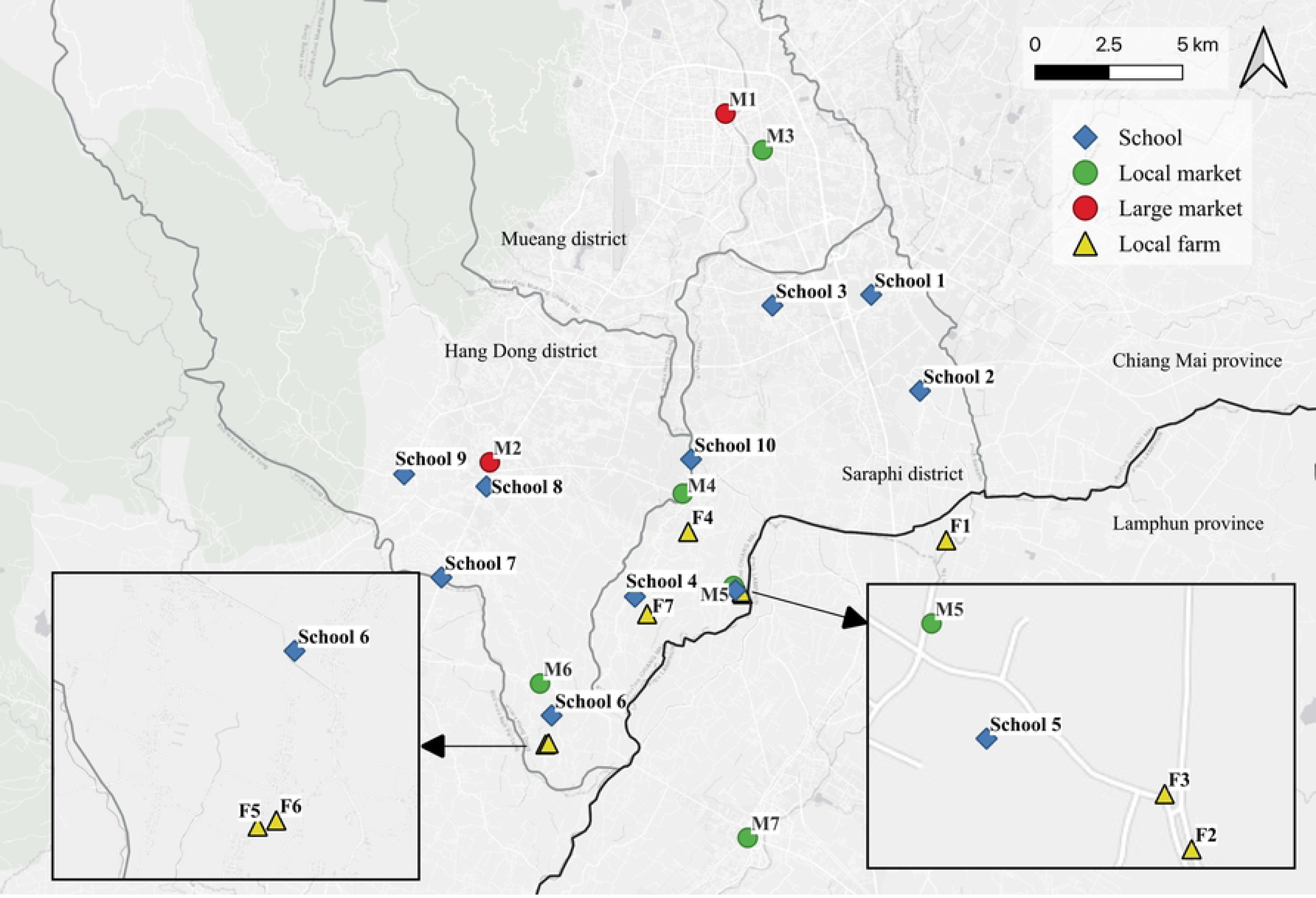
Locations of schools, markets, and local farms in the Saraphi and Hang Dong districts of Chiang Mai, Thailand. M1 = Mueang Mai Market; M2 = Hang Dong Market; M3 = San Pa Koi Market; M4 = Nam Tong Market; M5 = Local market (no official name); M6 = Ton Chok Market; M7 = Nong Dok Market; F1 = Farmer 1; F2 = Farmer 2; F3 = Farmer 3; F4 = Farmer 4; F5 = Farmer 5; F6 = Farmer 6; F7 = Farmer 7.

### Data collection

The 19 semi-structured in-depth interviews were conducted in Thai language, each lasting between 30 and 45 minutes. Interviews were conducted from August to September 2022, at school and farm locations. Interviews were recorded and later transcribed, and additional field notes were taken.

This study prioritized vegetables and fruits within the school lunch program due to their significant nutritional value and the persistent challenge of promoting sufficient consumption among children. Vegetables and fruits play a critical role in ensuring a balanced diet, preventing chronic diseases, and addressing micronutrient deficiencies in children [20–22]. In addition, sourcing vegetables and fruits from local farms in Thailand is relatively straightforward, offering schools an opportunity to support sustainable agricultural practices while enhancing the nutritional quality of school meals [23]. While the definitions of “local” vary, ranging from food grown within 100 km to that produced within the same state [24, 25], this study defines local food as produce grown or sourced within the same district as the schools. This practical framing aligns with how procurement and community engagement are organized at the local level.

Interview questions, prompts, and guides were provided by one of the authors (PP). The SWOT analysis questions were designed to comprehensively assess the feasibility of integrating local farms into the school lunch program. As summarized in Table 1, the focus areas included internal factors, such as current school lunch practices, procurement approaches, and farm production capacity, as well as external conditions, including support mechanisms, policy constraints, and market challenges. Questions for school staff addressed the current structure of their lunch programs, procurement practices, challenges encountered, and perceived opportunities for improvement. For farmers, the questions centered on their production methods, market access, challenges, and potential opportunities for collaboration with schools. At the time of data collection, 1 Thai baht was approximately 0.03 US Dollar.

**Table 1.** Key focus areas used in the swot analysis of school-farm integration in the Saraphi and Hang Dong districts of Chiang Mai, Thailand.

| **SWOT category** | **Focus area** | **School staff perspective** | **Farmer perspective** |
| --- | --- | --- | --- |
| Strengths | Program structure and capacity | Types of school lunch programs; current procurement practices; internal collaboration potential | Crop types, planning systems, customer base, readiness to supply schools |
|  | Feasibility of collaboration | Menu planning flexibility; coordination with farmers | Willingness to collaborate; required steps for school engagement |
| Weaknesses | Operational and logistical barriers | Unresolved lunch program issues; difficulties with direct farm sourcing | Challenges in direct school sales; farm sustainability issues |
| Opportunities | Potential for local integration | Willingness to buy from local farms; perceived long-term support needed | Potential market expansion; production planning flexibility |
| Threats | External barriers and risks | Threats to program; barriers to farm collaboration | External sales threats; logistical support needed; anticipated school collaboration challenges |

### Data analysis

In this study, the thematic analysis approach, which is widely used for identifying, analyzing, and reporting patterns or themes within data [26], was applied for the data analysis. Thematic analysis was chosen for its flexibility and ability to reveal detailed insights, making it particularly effective for understanding complex issues [27]. This approach enabled the identification of key facilitators and barriers influencing local food sourcing in Thai school lunch programs.

Interview data were analyzed manually using Microsoft Excel, following an inductive thematic analysis approach as outlined by Naeem [28]. Even though there are Computer-Assisted Qualitative Data Analysis Software (CAQDAS) programs that support Thai script, they often struggle with automatic word segmentation and keyword queries due to the structure of the Thai language, which lacks explicit word boundaries. Therefore, manual coding was preferred in this study to ensure accurate interpretation of the data. In the initial step, all interview data were transcribed word-for-word in Thai. Relevant quotes from interviews and field notes were selected and translated into English. Keywords were identified from the data and organized into codes based on recurring patterns. These codes were subsequently developed into themes that captured the key ideas emerging from the data.

To further deepen the analysis, a framework analysis was conducted using the SWOT framework. This allowed for a structured examination of the factors influencing the feasibility of integrating local farms into the school lunch program, echoing the approach taken by Skinner et al. [29] in their study of a First Nations community. Categorizing themes into the four SWOT quadrants enabled a comprehensive analysis of the internal and external factors influencing school-farm integration. In the context of this study, internal factors refer to any resources, constraints, or capacities under the joint control of schools and local farms. By contrast, external factors encompass conditions beyond their direct influence. This distinction ensures that strengths and weaknesses focus on what schools and farms can realistically manage internally, while opportunities and threats reflect influences from outside that partnership’s immediate scope.

The TOWS matrix, a strategic planning tool, was then employed to develop actionable recommendations based on the identified themes. This matrix systematically analyses the interplay between internal and external factors to generate strategic options [16, 29]. The TOWS matrix builds upon the SWOT analysis by providing a structured framework for developing strategies that leverage strengths to capitalize on opportunities (SO), utilize strengths to mitigate threats (ST), overcome weaknesses by exploiting opportunities (WO), and minimize weaknesses to avoid threats (WT). This approach ensures that the strategic recommendations are grounded in a thorough understanding of the internal and external contexts influencing the integration of local farms into the school lunch program.

Two additional researchers independently reviewed and verified the coding and analysis process to ensure the accuracy, consistency, and reliability of the findings. This inter-rater reliability check aimed to minimize potential biases and enhance the trustworthiness of the qualitative data analysis.

## Ethical approval

Ethical approval for this study was granted by the Centre for Development Research (ZEF) at the University of Bonn, Germany, on July 8, 2022. The approval documents were translated into Thai and submitted to the officers at Chiang Mai Primary Educational Service Area Office 4. Permission was then sent to contact the schools. Information sheets and consent forms were provided to school staff and local farmers, and the signed forms were returned to the researchers prior to participation.

## Results

### Participant characteristics

Table 2 provides an overview of the schools’ characteristics. In most schools, lunch ingredients were sourced through daily or weekly market visits by school staff. Some schools outsourced meal preparation to external vendors, with limited use of formal procurement agreements. School 5 had a different lunch service model. Due to its limited budget and staff, the school forwarded the lunch budget to parents, and students brought lunch from home. Table 3 presents the demographic information of the key informants. Participants included six local farmers and 13 school staff members, who were primarily responsible for managing the school lunch: directors (n=5), lunch teachers (n=5), and cooks (n=3).

**Table 2.** School characteristics in the Saraphi and Hang Dong districts of Chiang Mai, Thailand.

| School | District | Enrolment<sup>a</sup> | Lunch service | Main procurement method |
| --- | --- | --- | --- | --- |
| School 1 | Saraphi | 209 | Hiring a cook | Market (San Pa Koi) |
| School 2 | Saraphi | 132 | Contracting with<br>a lunch provider | Market (Mueang Mai) |

| **School** | **District** | **Enrolment<sup>a</sup>** | **Lunch service** | **Main procurement method** |
| --- | --- | --- | --- | --- |
| School 3 | Saraphi | 260 | Hiring a cook | Market (Mueang Mai) |
| School 4 | Saraphi | 85 | Hiring a cook | Local farm and market (Nam Tong) |
| School 5 | Saraphi | 68 | Bringing lunch from home | Market (Nong Dok) |
| School 6 | Hang Dong | 81 | Hiring a cook | Market (Ton Chok) |
| School 7 | Hang Dong | 205 | Contracting with a lunch provider | Market (Hang Dong) |
| School 8 | Hang Dong | 198 | Contracting with a lunch provider | Market (Hang Dong) |
| School 9 | Hang Dong | 350 | Contracting with a lunch provider | Market (Mueang Mai) |
| School 10 | Hang Dong | 131 | Hiring a cook | Market and local farm |

<sup>a</sup>Number of students from kindergarten to 6th grade

**Table 3.** Characteristics of participants in the Saraphi and Hang Dong districts of Chiang Mai, Thailand.

| Variables | Number of school staff | Number of farmers |
| --- | --- | --- |
| **District** |  |  |
| Saraphi | 7 | 3 |
| Hang Dong | 6 | 3 |
| **Age (years), median (P25, P75)** | 50.9 (44.5, 60.0) | 66.6 (56.5, 68.7) |
| **Gender** |  |  |
| Male | 3 | 3 |
| Female | 10 | 3 |
| **Highest education, n (%)** |  |  |
| Lower than 9<sup>th</sup> grade | 1 | 3 |
| 12<sup>th</sup> grade/vocational | 2 | 0 |
| Bachelor's and above | 10 | 3 |
| **Work experience** |  |  |
| 1-10 years | 8 | 3 |
| 11-20 years | 3 | 0 |
| >20 years | 2 | 3 |

Information was also gathered on, and observations were made at the markets where school staff purchased ingredients for school lunches. Table 4 summarizes the key characteristics of these markets, including their scale, product offerings, and location features.

**Table 4.** Market characteristics (based on observation) in the Saraphi and Hang Dong districts of Chiang Mai, Thailand.

| Market name | Market type | Key characteristics |
| --- | --- | --- |
| Mueang Mai (M1) | Large | A wholesale market in Chiang Mai city; features a large building with numerous vendors and a wide variety of goods |
| Hang Dong (M2) | Large | A large-scale in Hang Dong district; serves a substantial community; offers a wide variety of goods |
| San Pa Koi (M3) | Local | A community-based market in Chiang Mai city, focuses on fresh produce and street food; known for its local favourites |
| Nam Tong (M4) | Local | A market in Hang Dong district; serves a large community; primarily features local produce. |
| Local market (M5)<br>(No official name) | Local | A small-scale in Saraphi district; focuses on local products |
| Ton Chok (M6) | Local | A market in Hang Dong district; focuses on local products |
| Nong Dok (M7) | Local | A community-based market in a less densely populated area of Lamphun province |

**Fig 2** shows the distances (km) between schools, markets, and local farms, with distances measured from each school to its primary ingredient procurement location. The observed longest distance was 20.9 km from School 9 to Mueang Mai market (M1). Only two schools, i.e., School 4 and School 10, sourced fruits and/or vegetables from local farms. The cook at School 4 primarily sourced fruits and vegetables from Farmer 7. However, during the longan (*Dimocarpus longan* Lour. [Sapindaceae]) harvesting season in September, when Farmer 7’s production is limited to longan, the cook at School 4 bought ingredients from Nam Tong Market (M4). For School 5, staff previously purchased ingredients at the Nong Dok Market (M7). A small local market (M5) and two local farms (F2 and F3) were also located in the vicinity of this school.

**Fig 2.**
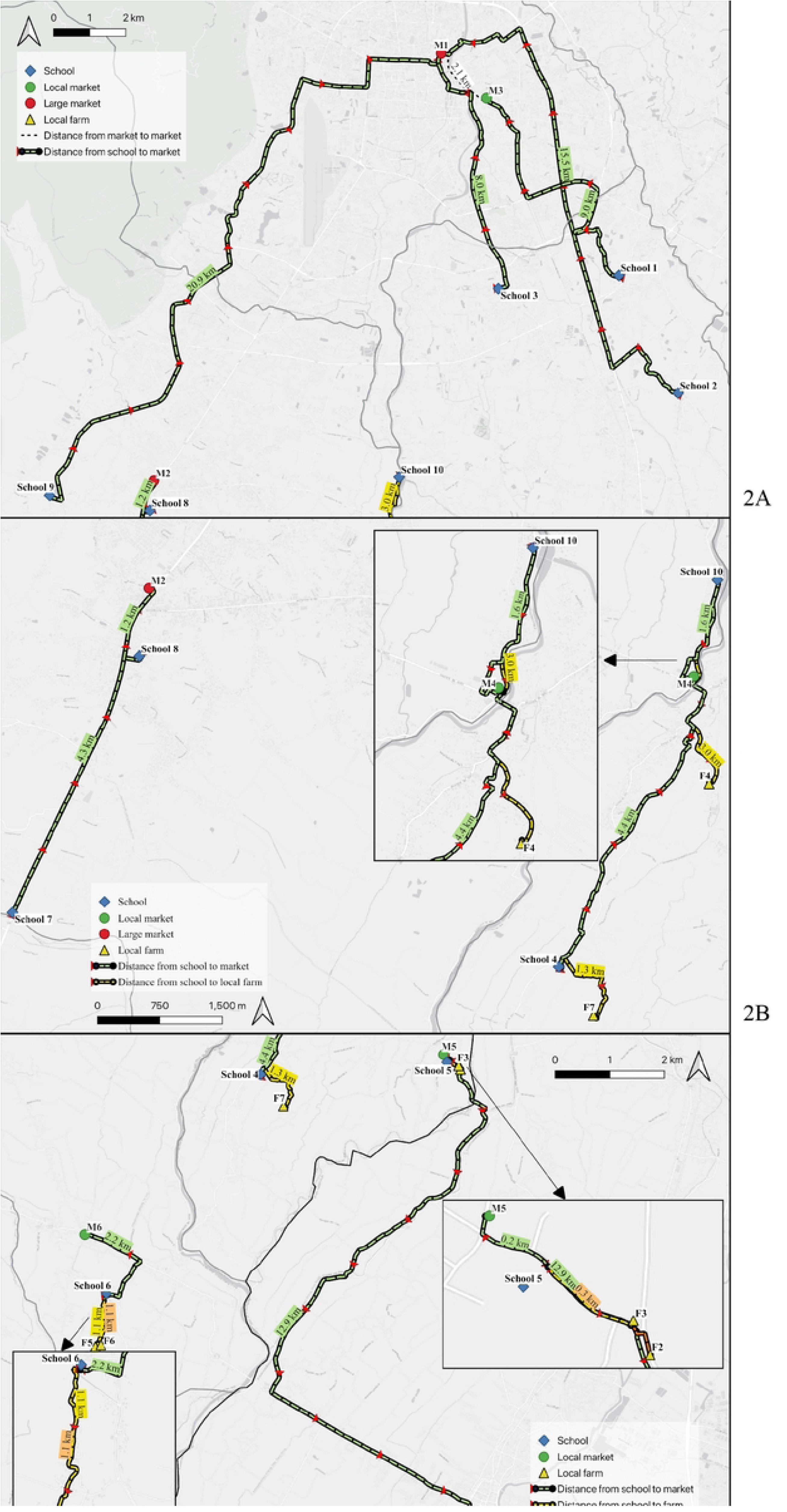
Distance (km) from schools to their primary ingredient procurement locations in the Saraphi and Hang Dong districts of Chiang Mai, Thailand. 2A: Distances from Schools 1, 2, 3, and 9; 2B: Distances from Schools 4, 7, 8, and 10; 2C: Distances from Schools 5 and 6.

Among the farmers, one participant (Farmer 6) cultivated land that he did not own. The land belonged to a landowner who lived in another city and offered its use in exchange for informal caretaking. Although the landowner did not maintain the perennial fruit trees, Farmer 6 grew seasonal vegetables and raised animals on the plot. All income generated from selling the produce remained with the farmer, even though they did not own the land.

### SWOT findings

These findings respond to the SWOT key areas (Table 1), which guided the thematic structure of the interviews. Internal factors (strengths and weaknesses) were drawn from school and farm practices under their control, while external factors (opportunities and threats) reflected wider contextual influences beyond their direct authority.

### Strengths within the school-farm system

In total, six internal strengths within the Thai school lunch program and local farms could be identified: robust community support, efficient school meal management practices, well-established procurement strategies, supportive nutritional environments, agricultural education initiatives, and adequate infrastructure. Collectively, these strengths underpin viable partnerships aimed at enhancing local food sourcing.

### S1: Community support in school food and agriculture

A prominent strength highlighted by the interview data was the community engagement that underpins school lunch programs. School staff indicated that their schools had strong relationships with local farmers, who often offer lower-cost produce directly to schools. The cook from School 4 shared, *“I buy vegetables from a local farmer because they (vegetables) are cheaper than the market. Some people grow vegetables at home, and they deliver them to the school.”*, emphasizing how farmers can provide cost savings while reducing the time teachers or cooks spend travelling to markets.

Participants reported the availability of organic or pesticide-free vegetables at competitive prices when purchased directly from local farms. *“Sometimes, I buy from an organic farm…it is cheaper than the market, which is 10-15 baht more expensive,”* said the cook from School 4. These examples highlight how direct relationships with farmers help schools predict supply and cost more reliably.

Moreover, some cooks lived within the school’s neighborhood. For example, *“The cook’s house is located across from the school, and she lives in our community,”* shared the director of School 7. This type of proximity and familiarity appeared to support informal coordination and ease of daily meal preparation. Similarly, community ties extended beyond the school itself. This is reinforced by Farmer 1, who shared, *“Teachers from School 2 said they would bring the children to visit our farm and also cook food here*.*”*

### S2: Efficient and structured school lunch management

Another major strength, repeatedly underscored by interviewees, was school-level organization and clarity of responsibilities. Participants described systems where teachers, principals, and contracted cooks operate under well-defined guidelines, with some reporting few challenges in day-to-day operations. As the director of School 7 put it, *“I have used the same system since 2020…it has been running well.”* By consistently applying this system, covering everything from menu creation to budget monitoring, schools maintain stable, reliable lunch programs.

Accountability measures within school lunch programs emerged as significant. Schools commonly retained the right to withhold payment if cooks deviated from standards and regularly conducted meal quality checks using photographs or sign-off sheets (Director, School 7). These quality control measures not only ensured compliance with national nutrition guidelines but also enabled schools to communicate expectations to suppliers, including local farmers.

Additionally, participants emphasized the strength of detailed menu planning and nutritional oversight. Following the Thai school lunch standard, menus are often rotated to ensure variety. One participant elaborated on a system where fruit or dessert was provided daily, fostering balance in children’s diets (Director, School 7; Lunch Teacher, School 8). Another participant explained that the teacher-cook collaboration ensured alignment with guidelines while adapting to seasonal vegetables and fruits. *“We adapt the menu based on what fruits and vegetables are available in that season”* (Cook, School 2).

### S3: Procurement strategies and local food sourcing

Interview data revealed an array of procurement strategies to secure fresh ingredients. Participants reported frequent market visits, with teachers or cooks taking turns to shop based on personal schedules (Cook, School 2; Director, School 6). Meanwhile, schools closer to a large wholesale supermarket combined these sources with local produce, optimizing cost and variety. One participant summarized: *“I buy vegetables and meat every day…We also cultivated some mushrooms with the children’s help”* (Cook School 2), illustrating the integration of daily shopping, local sourcing, and occasional self-production.

Seasonality emerged as a key factor, as certain crops, like bananas or cabbage, became more abundant and cheaper during specific times of the year (Cook, School 2). By anticipating price fluctuations and seasonal availability, staff could make cost-effective choices while sustaining variety in meals. While external markets remained a primary avenue, as shown in Table 2, local farms consistently offered a viable and sometimes preferable alternative when the right conditions, such as consistent supply and aligned planting schedules, were met.

### S4: School food environment and student nutrition management

Alongside procurement, schools implemented strong internal controls over their food environments to promote healthy eating. This included regulating snack vendors and encouraging healthier in-school options. *“We stopped external vendors from entering the school. It was part of our policy,”* shared the director of School 7. In other cases, schools encouraged the use of healthier cooking methods: “*We switched from frying to steaming snacks,”* said the same director.

While not all these points directly touched on local farm produce, they underscored an ethos of prioritizing children’s health. By creating a controlled food environment, schools laid the groundwork for deeper partnerships with farmers who supply fresh, nutrient-dense items.

### S5: School garden and agricultural education

The integration of school gardens with learning activities emerged as another strength. School staff explained how teachers, students, and sometimes community members worked together to grow vegetables like eggplant and water spinach, or to maintain a small rice plot (Director, School 7; Director, School 8). Some schools even engaged in mushroom cultivation, compost making, and integrated farming activities. The director of School 8 recounted, *“We grow water spinach…use it for school lunches…even sell some to the parents.”* These examples indicated that school gardens could deliver multiple benefits: reducing procurement costs, providing a hands-on learning platform, and reinforcing the idea that fresh, local produce was both accessible and valuable. The gardens reduced procurement costs and offered students practical experience in agriculture and nutrition, strengthening the foundation for future farm-to-school engagement.

This school-based approach to agricultural learning also aligned with farmers’ perspectives. As one local farmer shared, *“We do this to help the villagers by providing training on various topics…we grow mushrooms, make fertilizer, basically do everything ourselves”* (Farmer 2). Such synergies between community farming initiatives and school garden programs highlight a shared commitment to agricultural education and strengthen the potential for long-term school–farm partnerships.

### S6: School infrastructure and resource management

Infrastructure was found to be a facilitating factor. One school director described how having two buses enabled convenient trips to farms and agricultural learning centers, without incurring additional travel costs (Director, School 4). Although the buses were not typically used for ingredient procurement, they supported student visits that reinforced agricultural learning and awareness of local food systems. Teachers and cooks could also store fresh produce longer due to access to refrigeration, supporting larger, less frequent shopping trips. These resources are aligned with Table 1’s questions regarding feasibility and logistical coordination. When schools are equipped to store, transport, and manage larger quantities of perishable goods, they are better positioned to collaborate with local producers. This logistical capacity, coupled with clear administrative systems, made some schools well-prepared to scale up farm-to-school initiatives.

### Weaknesses within the school-farm system

The weaknesses highlighted internal limitations faced by the Thai school lunch program practices and associated local farms. These encompass budgetary constraints, procurement difficulties, insufficient oversight, inadequate facilities, and challenges aligning menus with student preferences. Such factors significantly impede the effective integration of locally sourced produce into school meals.

### W1: Budget constraints and financial management

School staff repeatedly emphasized that the statutory allocation of 21 baht per student per day limited their ability to provide meals that were both varied and nutritionally adequate. In one case, a school returned the lunch budget to parents in cash and asked them to prepare lunches for children. A teacher explained, *“I contacted a vendor to provide school meals at 19-20 baht per meal. The total budget was 21 baht per meal, so I asked if we could cut 2 baht to buy fruit or snacks for the children. They agreed, but ingredient prices kept increasing. So we dropped this plan.”* (Lunch Teacher, School 5). Additionally, rising food prices intensified financial strain, making it more challenging to consider higher-quality or locally grown options. These responses highlighted the financial fragility of the program and its dependence on low-cost ingredients.

### W2: Procurement strategies and local food sourcing

Despite interest in direct sourcing, many schools faced logistical hurdles, such as seasonal constraints, limited local agricultural output, and transportation issues that restricted reliable farm-to-school integration. One teacher shared, *“If we grow vegetables ourselves…the quantity is not enough…if we need a large quantity, we must buy from markets like Mueang Mai”* (Lunch Teacher, School 8). These responses reflected the difficulty of aligning school procurement needs with what local farms could consistently supply, addressing challenges in purchasing directly from farms and unresolved procurement problems.

### W3: School food environment and nutrition management

Although some schools fostered a healthier on-campus food environment, limited staffing and resources made it challenging to monitor children’s snack choices. A participant noted that *“The students eat all three times…they might not eat much for lunch but prefer snacks, beverages, and sugary drinks…we do not know how to solve this issue”* (Lunch Teacher, School 9). Without sufficient oversight or funding to promote healthier alternatives, schools struggled to ensure that local, high-quality produce was prioritized over cheaper, less nutritious snacks.

### W4: School garden and agricultural education

Many schools recognized the value of on-campus gardens in supplementing lunch ingredients and teaching students about agriculture. However, maintenance remained a persistent issue, particularly when teacher availability or funding was insufficient. Some gardens offered only minor yields, limiting their contribution to daily meals. In other instances, the COVID-19 pandemic led to staffing shortages that stalled garden activities altogether. For instance, one school experienced rising costs of chicken feed in their garden-based poultry project, remarking, *“At first, we fed them with regular chicken feed, but it was expensive, and they rarely laid eggs…they have become quite plump”* (Director, School 7). As a result, these gardens could not consistently supply the volume needed for full integration into school lunches.

These limitations directly address the SWOT question concerning unresolved challenges and the feasibility of garden-based food production. Moreover, the difficulties experienced in school settings were echoed by farmers. For example, Farmer 6, who cultivated land offered by a non-resident landowner, noted challenges with tree maintenance*: “Guavas produce fruit all year round…but they lack proper care. The landowner does not maintain the farm.”* In this case, the landowner allowed use of the land but did not contribute to its upkeep, leaving the farmer responsible for all maintenance. Although he received all income from the produce, managing everything independently required significant time and effort, which sometimes made it difficult to maintain consistent productivity.

### W5: Infrastructure and resource limitations

In relation to logistical and physical constraints, participants described challenges with inadequate kitchens, storage areas, and cooking spaces. One school had to outsource meal preparation because the cook did not have access to an on-site kitchen: *“The cook prepares the meals at home and then delivers them to the school because there is no cafeteria here”* (Director, School 7). These deficiencies, mentioned in response to sourcing and feasibility questions, restricted schools’ ability to store or process farm-fresh produce efficiently, reducing the feasibility of farm-to-school integration.

### W6: Lunch management and alignment with guidelines

Staff discussed varying degrees of menu planning, record-keeping, and compliance with the Thai School Lunch Standards issued by the Ministry of Education. In some instances, the school contracted meal preparation out to cooks with minimal supervision, while children often discarded fruits or vegetables they disliked. The lunch teacher of School 1 explained that *“We hired a cook on a contract basis, and we do not get involved at all. She handles everything herself…”* This detachment reduced accountability, leading to waste when students rejected unfamiliar fruits and vegetables. When asked about menu design and guideline alignment, participants noted the difficulty of enforcing nutritional standards under current contracts and budget constraints.

### Opportunities for school-farm integration

Opportunities identified through the interview data revealed promising avenues for future collaboration, especially in leveraging community resources, simplifying procurement, and strengthening agricultural education.

### O1: Community support in school food and agriculture

Participants reported that existing relationships with villagers and local authorities provided tangible assistance to school food provision and agricultural activities. One director mentioned that villagers willingly donated unused land for rice cultivation, noting, *“This is not school property; it belongs to the local villagers…whatever rice is harvested, they donate all of it to the school without asking for any payment”* (Director, School 7). This points to the potential for community-driven solutions, particularly in areas with strong social cohesion. In addition, existing municipal support, such as organizing field trips, could be strategically expanded to integrate educational visits, produce delivery, or farm-based learning into school lunch programming. A local farmer reinforced this potential, recalling, *“The municipality partnered with a private company to help prepare the soil for us. After that, people could plant whatever they wanted”* (Farmer 1). These examples illustrate how participants perceived support from both community members and local authorities in facilitating food production or education-related activities.

### O2: Procurement strategies and local food sourcing

Participants noted that fruit and vegetables were often purchased informally from neighboring farmers, village vendors, and household orchards. One cook mentioned, *“If it is available locally, we use what is available. It is better than going out to buy from the Mueang Mai Market or dealing with chemicals…”* (Cook, School 2). School staff believed that shorter supply chains lowered travel costs and delivered fresher produce. They suggested that these informal links could be formalized into regular supply agreements, thereby ensuring consistent volumes and prices. A school director said: *“Like guavas…I know a farm near my home…I bring [jackfruits] from my home for students”* (Director, School 10). Participants therefore viewed written purchase schedules and harvest partnerships with local farmers as a practical way to align menus with seasonal availability while strengthening ties between schools and their surrounding farmers.

### O3: School garden and agricultural education

School gardens and agricultural education emerged as a promising avenue for deepening farm-to-school collaboration. Some participants described teaching programs or visits to government-supported agricultural learning centers, which are demonstration farms that showcase practices in sustainable, low-input agriculture, such as *“taking students to meet local wisdom experts is part of a learning reduction program…We also take them to a government-supported agricultural learning center for a one-day trip”* (Lunch Teacher, School 5). These efforts promote an agricultural mindset among teachers and students, cultivating an environment more receptive to the integration of locally produced food. Expanded school gardens, composting stations, and student-managed plots were viewed as opportunities to enhance school menus while reducing expenditures on vegetables.

### Threats to school-farm integration

Responses to questions on external threats, obstacles, and challenges in local procurement revealed three major issues: funding instability, seasonal variability in supply, and limited farm capacity, all of which can disrupt or delay the adoption of farm-to-school initiatives.

### T1: Rising costs and inflation

School staff identified inflation and the rising cost of food as major external threats. Participants noted that essential ingredients such as pork, eggs, and vegetables had become increasingly expensive, further straining the already limited 21-baht per student per day lunch budget. The lunch teacher from School 5 highlighted the impact, stating, *“Right now, prices have increased significantly. Pork has gone up to almost 200 baht per kilogram, and today eggs have increased to 5 baht per egg”*. These economic pressures reduced schools’ ability to procure high-quality or locally sourced ingredients and forced compromises in both nutritional variety and food quantity. Unlike internal budgetary constraints (W1), which are static and institutionally defined, T1 reflects dynamic and unpredictable economic conditions that amplify existing vulnerabilities. This economic strain is not confined to schools alone. Farmers also reported facing rising input costs, particularly for fertilizers. *“Fertilizer that used to cost 900 or 1,000 baht is now 1,700 to 1,900 baht,”* noted Farmer 4. This shared burden across schools and farms further complicates efforts to establish reliable, cost-effective farm-to-school linkages.

### T2: Community support inconsistency

While some communities actively shared surplus produce, others paused growing vegetables due to seasonal harvests or labor constraints, limiting consistent sourcing. One teacher explained that “*People are not growing them [vegetables] because it is the longan season…they are busy harvesting longan, so vegetables are not being harvested”* (Lunch Teacher, School 4). In other areas, most residents worked in construction or city-based jobs, leaving minimal time for farming. These fluctuations in community participation pose a challenge to the sustainability of school-farm partnerships, as schools cannot reliably depend on local fields to provide produce throughout the academic year.

### T3: Local supply shortfalls

Participants highlighted persistent challenges related to the limited capacity of local farms to meet school needs. Many schools required a steady volume and variety of ingredients, yet local farms were unable to provide consistent or large-scale output. For instance, when sourcing vegetables, some schools reported purchasing high volumes daily, a demand that far exceeded what most smallholders could supply. One cook remarked that *“The local community market does not have enough produce for our school…we buy approximately over 20 kilograms per day”* (Cook, School 8). As a result, schools often defaulted to large markets like Mueang Mai, where produce was more reliably available in bulk. This structural mismatch between school needs and community-scale production created a dependency on centralized suppliers, which undermined efforts to localize procurement and weakened the resilience of school-farm integration.

### TOWS analysis and strategic recommendations

The TOWS analysis expands upon the SWOT findings by developing concrete, context-specific strategies to address the second research question: What strategies can enhance the linkage between Thai school lunch programs and local farms? By aligning internal factors with external conditions, the analysis offers practical recommendations that schools, farmers, and policymakers can implement to strengthen farm-to-school integration. Fig 3 illustrates how strengths and weaknesses within schools and farms interact with external opportunities and threats, guiding the identification of context-specific strategies.

**Fig 3.**
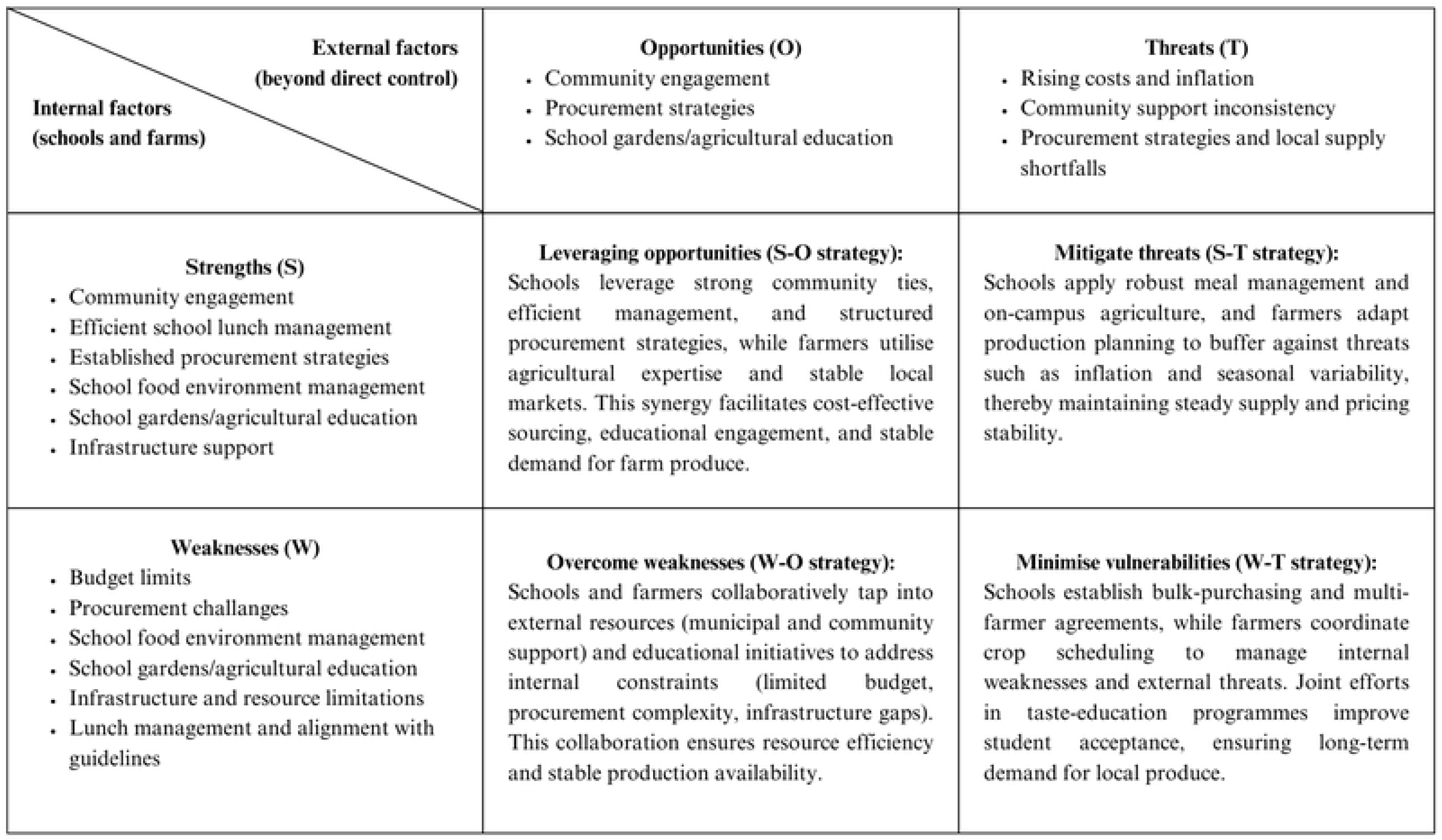
Conceptual model of school-farm collaboration in the Thai school lunch programme in the Saraphi and Hang Dong districts of Chiang Mai, Thailand.

### S-O (strengths + opportunities)

This strategy focuses on using existing school and community strengths to take full advantage of external opportunities. Schools with strong ties to local farmers (S1) can formalize formal/informal agreements into structured supply partnerships, taking advantage of community goodwill and municipal support (O1). For example, schools may draft procurement contracts or harvest calendars with farmers to secure regular seasonal produce, reducing costs and increasing predictability.

Schools with established procurement systems (S3) and proactive food environment management (S4) can pair these strengths with local farm partnerships (O2) to diversify supply chains and reduce reliance on distant markets. Concrete actions include assigning a procurement lead to coordinate with farmers on seasonal availability and aligning school menus to integrate these ingredients in ways that promote student acceptance, such as themed fruit days or local harvest menus.

Moreover, infrastructure such as school buses and refrigerators (S6) support both logistical and educational activities. While school buses are primarily used for student transport and farm-based learning trips, such visits can help strengthen school-farm relationships and deepen agricultural understanding (O3). In parallel, access to refrigeration allows schools to store perishable goods, making it easier to incorporate locally sourced produce when available even if deliveries occur only a few times per week.

### S-T (strengths + threats)

In S-T strategies, schools use internal strengths to defend against or mitigate external threats. Even though local farming cycles are irregular or ingredient prices surge, schools that have reliable administrative systems, robust school gardens, or supportive communities can withstand these challenges more effectively. This pairing ensures that the positive internal factors offset risks posed by inflation, seasonal supply shortages, or other market uncertainties.

For instance, schools with efficient lunch management systems (S2) can mitigate the effects of inflation (T1) by employing structured menu planning, locking in bulk or seasonal pricing, and adjusting ingredients without compromising nutritional quality. Similarly, reliance on school-grown produce (S4) offers a buffer during periods of seasonal scarcity (T2), ensuring a consistent, if limited, supply of fresh vegetables and herbs.

Furthermore, schools with established procurement structures and storage infrastructure (S6) can rotate menus based on available produce and maintain consistent inclusion of fruits and vegetables, reducing dependence on centralized markets (T3). As local farmers observe steady demand from schools, they may be encouraged to adjust their planting schedules, further supporting localized, sustainable supply chains over time.

### W-O (weaknesses + opportunities)

Schools address or overcome internal weaknesses by taking advantage of external opportunities. Budget constraints, staff shortages, and infrastructural inadequacies can be mitigated through community partnerships, municipal support, or the application of local knowledge and resources. By aligning school-level challenges with supportive conditions beyond their direct control, these weaknesses may be transformed into manageable, and in some cases, productive solutions

For example, budget constraints (W1) can be alleviated through collaboration with local authorities or temples. Some schools reported receiving donated rice or vegetables, which reduces expenditure and creates room for higher-quality or locally sourced ingredients (O1). Similarly, procurement-related burdens (W2) and staff availability issues (W3) can be reduced through formal/informal agreements with nearby farmers or orchard owners (O2), who can deliver produce directly to schools. Such arrangements limit the need for daily market visits, ensure a more consistent supply, and strengthen local relationships.

Besides, shortcomings in infrastructure or equipment (W4, W5) can be addressed through support from government organizations or collaborative projects tied to agricultural education (O3). This includes improvements to kitchen facilities, storage areas, or garden space, enabling schools to more effectively store, process, or grow fresh produce. In turn, these improvements contribute to both nutrition and learning, reinforcing the educational dimension of farm-to-school initiatives.

### W-T (weaknesses + threats)

W-T strategies focus on the most vulnerable intersection, where internal shortcomings face external pressures. Schools experiencing budget shortfalls, inconsistent oversight, or limited staff capacity face additional challenges from rising inflation and the seasonal inconsistency of local farm produce. This quadrant focuses on strategies that minimize or adapt to such compounding difficulties, ensuring that schools can continue to provide nutritious meals and sustain partnerships with local farms under adverse conditions.

Financial constraints (W1) compounded by inflation (T1) necessitate proactive cost-management measures. These might include bulk purchasing agreements, the use of school-grown produce to supplement meals, or the renegotiation of fixed-term supply contracts to stabilize pricing. Similarly, procurement difficulties (W2) combined with seasonal shortages (T2) call for collaboration with multiple farmers, allowing staggered crop cycles to ensure continuous availability throughout the school term. In parallel, student reluctance to consume certain vegetables (W6) alongside dependency on centralized food sources (T3) risks increased waste and a fallback on ultra-processed alternatives. To counter this, food literacy initiatives such as taste-testing, cooking demonstrations, or “farm-to-lunch” days can improve acceptance of local and seasonal ingredients, supporting a more sustainable and locally integrated school lunch program.

## Discussion

This study aimed to explore the drivers and barriers influencing collaboration between the Thai school lunch program and local farms and to identify strategies that enhance this integration. The findings from the SWOT-TOWS analysis provided detailed insights into the key factors that shape successful partnerships, along with specific obstacles that hinder these collaborations.

Regarding the drivers, community engagement emerged prominently as an internal strength, providing essential support that facilitated direct procurement from local farms. This finding aligns with previous studies by Joshi et al. [10] and Izumi et al. [17], which emphasized the pivotal role of community partnerships in the success of farm-to-school programs. However, this study uniquely illustrates the practical implications of community engagement in semi-urban contexts, such as those found in Saraphi and Hang Dong districts of Chiang Mai. Unlike purely urban or rural settings explored in prior research [30, 31], the semi-urban environment in Thailand presented distinctive dynamics where community ties significantly facilitated access to affordable and local produce.

Efficient school meal management practices also proved critical in underpinning robust collaboration. Clear organizational roles, accountability through regular quality checks, and transparent procurement processes were integral internal strengths. These findings corroborate the broader literature, emphasizing structured management as essential for sustaining farm-to-school initiatives [32]. In addition, this study provides fresh insights by highlighting specific accountability mechanisms, such as photographic documentation and daily sign-off sheets, practices not extensively documented in previous research. These approaches provided local farmers with clearly defined expectations, fostering reliability and trust, essential components of sustainable partnerships.

Barriers identified included budgetary constraints, logistical challenges related to seasonality, and inadequate infrastructure. Financial limitations, reflected in the allowance of only 21 baht per student per day, severely restricted menu diversity and forced schools to procure ingredients from distant, centralized markets instead of local farms. Similar budget constraints have been noted elsewhere as major impediments to sustainable school food systems [30]. Yet, the distinct contribution of this study lies in quantifying and contextualizing these constraints within the Thai semi-urban school system, providing targeted insights for policymakers.

Infrastructural inadequacies, such as limited refrigeration and outdated kitchen facilities, posed additional barriers. These findings are consistent with prior research reporting infrastructure as a barrier to implementing farm-to-school programs [33]. The current study extends existing knowledge by demonstrating how these constraints specifically limit the handling and storage of fresh, locally-sourced produce, directly influencing procurement decisions.

External threats further complicated local sourcing, particularly due to the limited capacity of local farmers to produce the necessary volume and variety of vegetables and fruits. This resulted in reliance on centralized markets, undermining the goal of local sourcing. While prior studies broadly acknowledged issues with seasonal availability and production volumes [10, 17], this study uniquely delineates these challenges in the context of Chiang Mai’s farming capabilities, where farmers generally operated at smaller scales, affecting the sustainability and consistency of local produce supply.

Regarding policy implications, the study suggests that local sourcing can serve as a viable solution to enhance the sustainability and nutritional quality of school lunches, but this requires systemic support. Policymakers have a crucial role to play in promoting farm-to-school initiatives by providing targeted subsidies, training programs, and logistical assistance [34]. The findings indicate that policies fostering closer ties between schools and local farms, such as providing incentives for schools to purchase local produce, can help address the logistical and financial barriers identified in this study. This aligns with research on the effectiveness of procurement incentives in farm-to-school programs, which shows that per-meal reimbursement can increase local food purchases, especially when aligned with the growing season [35]. Moreover, creating partnerships with food hubs and local food distributors can help schools overcome the logistical challenges of sourcing from local farms [17].

This study revealed that the previous school lunch budget allocation of 21 baht per student per day significantly constrained schools’ capacity to implement sustainable local sourcing practices. Nevertheless, following the data collection for this study, the Thai government introduced a revised budget in the 2022 fiscal year, increasing the allocation to between 22 and 36 baht per student per day, depending on school size [36].

Despite these positive developments, there remains an absence of specific policy frameworks directly supporting school-farm collaborations. While the Thai government does provide limited funding for school garden initiatives, securing this funding typically requires additional administrative tasks, such as preparing detailed proposals, adding significantly to the existing workload of school staff. Previous literature highlights how administrative burdens can impede the successful implementation of farm-to-school initiatives by discouraging school staff from pursuing available funding opportunities [32, 34]. Therefore, to fully realize the potential benefits of local sourcing in school lunches, future policy initiatives in Thailand should aim to explicitly support and streamline collaborations between schools and local farms, alleviating administrative burdens and providing consistent logistical and financial backing.

One of the key strengths of this study is the valuable insights gained from local perspectives, including school staff and local farmers. While budget constraints are often a major challenge in school lunch programs, on-the-ground strategies that school staff employ to manage these constraints are crucially important. By focusing on local produce as a potential solution, the study sheds light on a viable option for improving the sustainability and quality of school meals. Finally, the employed qualitative design allows for an in-depth understanding of the operational challenges faced by schools, which quantitative methods may not fully capture.

While the study primarily considered perspectives of school staff and local farmers, future research could enrich understanding by incorporating views from other stakeholders, including students, parents, and policymakers. Investigating the nutritional impact of locally sourced produce could further demonstrate the practical benefits of local sourcing. Additionally, exploring the long-term financial sustainability and scalability of farm-to-school models in different contexts would be valuable. Lastly, addressing potential biases inherent in qualitative self-reported data by incorporating quantitative assessments would also strengthen future studies, providing more comprehensive insights into the efficacy of farm-to-school programs.

## Conclusion

The study highlights the potential drivers and barriers to integrating local farms into school lunch programs in Chiang Mai, Thailand. While locally sourced produce can enhance meal quality and promote community-based food systems, schools face persistent barriers, including seasonal variability, limited crop diversity, and financial and logistical constraints. Despite this, opportunities to strengthen school-farm collaboration through improved procurement planning, dedicated liaison roles, reduced administrative burdens, and enhanced municipal support do exist. To advance these collaborations, schools should improve menu planning and establish agreements with local farmers, while local authorities can provide supportive coordination, offering training, and investing in infrastructure. At the policy level, introducing procurement incentives, increasing lunch budget allocations, and simplifying funding access for school gardens are essential. Future research should investigate the implementation and impacts of these strategies in practice, particularly regarding food quality, cost-effectiveness, and student nutrition. Longitudinal studies evaluating health, educational, and economic outcomes would further provide insights into the sustainability and scalability of school-farm partnerships in the semi-urban contexts of Thailand.

## Data Availability

The data underlying this study contain potentially identifying or sensitive participant information and cannot be made publicly available to protect participant confidentiality. De-identified excerpts or summary data are available from the corresponding author (PP) upon reasonable request.

## Acknowledgements

We would like to thank Miss Piraporn Kiewkhen, who worked at the Creative Economy Agency (Public Organization) in Chiang Mai at the time of data collection, for her assistance in facilitating contact with the Office of Basic Education. We are also deeply grateful to all the participants who generously shared their time, insights, and experiences.

